# Efficacy of an Integrated Cognitive Control Training Program (ICCT) in Patients with Obsessive Compulsive Disorder: An Open-Label Trial

**DOI:** 10.64898/2025.12.23.25342773

**Authors:** Mahashweta Bhattacharya, Himani Kashyap, Srinivas Balachander, YC Janardhan Reddy

## Abstract

**Background:** Neuropsychological deficits are common in obsessive-compulsive disorder (OCD) and may influence functional and treatment outcomes. Only a few studies have effectively targeted these deficits, with most failing to show broad transfer of training. This study aimed to evaluate the efficacy of an integrated cognitive control training (ICCT) program on neuropsychological functioning in OCD patients and assess related changes in clinical and socio-occupational functions.

**Method:** A single-group open-label design with pre-, mid-, post-treatment, and follow-up assessments was employed with 38 participants diagnosed with OCD, who were on stable doses of serotonin reuptake inhibitors (SRIs). The ICCT program, integrating task- and game-based cognitive stimulation with metacognitive strategy training and generalization exercises, included 24 hours of training over eight weeks across therapist-guided and homework sessions. The intervention was systematically adapted and validated, and its efficacy was examined across neuropsychological, clinical, and socio-occupational domains.

**Results:** The intervention demonstrated moderate to large improvements in neuropsychological functioning (R²₍M₎ range = .15 to .27) and self-reported cognitive difficulties (R²₍M₎ = .58) and further demonstrated transfer to untrained domains such as OCD symptom reduction (R²₍M₎ = .54), anxiety (R²₍M₎ = .61), depression (R²₍M₎ = .60), metacognitive regulation (R²₍M₎ = .30), and socio-occupational functioning (R²₍M₎ = .26). However, response inhibition saw only small improvements (R²₍M₎ = .11).

**Conclusion:** The ICCT program achieved both near and far transfer of cognitive training, improving neuropsychological, clinical, and socio-occupational outcomes. This contrasts with prior interventions with limited transfer of training. The small effect on response inhibition may reflect the trait nature of the deficit, assessment limitations, or gaps in the intervention. Future studies should use randomized control designs to validate and compare ICCT with other interventions.

## Introduction

OCD is the fourth most prevalent mental disorder (Kessler et al., 2005), and is among the top 20 causes of illness-related disability for people aged 15 to 44 years (Markarian et al., 2010). Neuropsychological deficits are well-established in OCD, and most consistently evinced in planning, decision making, set shifting, inhibitory control, and visual memory (Abramovitch et al., 2013; Cavedini et al., 2010; N. Y. Shin et al., 2014). Recent research suggests that the neuropsychological deficits in OCD are caused by impairment in executive processes or cognitive control (Fruehauf et al., 2021; Kashyap et al., 2013; Snyder et al., 2015). Notably, these deficits do not fully respond to symptomatic treatment (Kim et al., 2002; Nielen & Boer, 2003; Riesel et al., 2015). A number of studies demonstrate that deficits in set-shifting, response inhibition (Bannon et al., 2006; Chamberlain et al., 2006; Olley et al., 2007; Rao et al., 2008), verbal fluency, planning, working memory (Kim et al., 2002), non-verbal memory (Kuelz et al., 2004) continue to exist during the remission phase (de Vries et al., 2019; Fruehauf et al., 2021) Importantly, some evidence suggests that neuropsychological deficits might predict pharmacotherapy and psychotherapy treatment outcomes in OCD (D’Alcante et al., 2012; Hybel et al., 2017; McNamara et al., 2014; Perna et al., 2016).

Cognitive training, also known as cognitive remediation, aims to improve cognitive deficits with transfer to underlying neural circuits and real-world functional abilities (Keshavan et al., 2014; Miyake & Friedman, 2012; B. A. Wilson, 2008). Originally developed for brain injuries, it has been applied to various psychiatric disorders like schizophrenia, attention deficit hyperactivity disorders, mood disorders, substance use disorders and anxiety disorders with varying levels of success (Bowie et al., 2013; Karbach & Kray, 2009; Kurtz, 2012; Sofuoglu et al., 2013). Despite the prominent and persistent neuropsychological deficits in OCD, cognitive training efforts are scanty. A recent systematic review of cognitive training in OCD found a total of only eight studies (Bhattacharya et al., 2024). These studies have shown mixed outcomes, with some reporting improvements in cognitive functions but only limited transfer to symptom severity or socio-occupational functions (Buhlmann et al., 2015; Calkins & Otto, 2013; Kashyap et al., 2019; Park et al., 2006; Rini et al., 2023).

Methodological factors possibly contributing to ‘far’ transfer (generalisation to untrained domains) have been extensively examined (Bhattacharya et al., 2024), in light of evidence-based recommendations for cognitive training. These include process-specific training focusing on variability of tasks and contexts (Slagter et al., 2011), adequate dose and duration of training (Lauenroth et al., 2016), and incorporating techniques of strategy monitoring (Cicerone et al., 2011; Tate et al., 2014). Three key components of training that have been identified are: Cognitive Activation, Strategy Planning, and Generalization, focusing on neuroplasticity, metacognitive strategies, and skill transfer (Bowie, 2019). A novel program previously tested in remitted OCD (Kashyap et al., 2019; Rini et al., 2023), has demonstrated transfer to untrained cognitive and clinical domains. In the present study, the Integrated Cognitive Control Training (ICCT) is examined in symptomatic OCD, with an objective to train cognitive control mechanisms in OCD and examine the potential impact on clinical outcomes and socio-occupational functioning.

## Materials and Methods

### Design

The study was conducted at the OCD Clinic and the Behavioural Medicine Unit of the National Institute of Mental Health and Neurosciences, Bengaluru, India, with approval from the Institute Ethics Committee (IEC; Behavioural Sciences, No. NIMH/Do/BEH. Sc. Div./2020-21). The trial was registered with the Clinical Trials Registry of India, ICMR, New Delhi (CTRI/2021/03/031770). The study adopted a single-arm, open-label design with baseline, post-assessment, and a three-month follow-up assessment.

### Outcome Measures

Primary outcomes included changes from baseline to post-intervention in self-reported cognitive difficulties (CAIOC-13) (Dittrich et al., 2011) and response inhibition, measured using the Stop Signal Task (Verbruggen & Logan, 2008).

Secondary outcomes included changes in OCD symptom severity, as measured by the Yale–Brown Obsessive Compulsive Scale—Symptom Severity (YBOCS) (Goodman et al., 1989); anxiety and depression, assessed using the Hamilton Anxiety Rating Scale (HAM-A) (Hamilton, 1959) and the Montgomery–Åsberg Depression Rating Scale (MADRS) (Montgomery & Asberg, 1979); metacognitive awareness and regulation, measured by the Metacognitive Awareness and Regulation Scale (MARS) (Vishwanathan et al., 2022); socio-occupational functioning, assessed using the Sheehan Disability Scale (SDS) (Sheehan et al., 2016); and neuropsychological performance across domains of attention, working memory, executive functions, and problem solving assessed using the Trail Making Test (Bowie & Harvey, 2006), Spatial Span (Gurappa, 2009), Controlled Oral Word Association Test (COWAT) (Benton et al., 1976), Zoo Map Test and Rule Shift Cards (B. Wilson et al., 1996), Complex Figure Test (CFT) (Osterrieth, 1944), and Block Design Test (Wechsler, 1997). An additional objectiv**e** was to examine the acceptability and feasibility of the intervention.

### Sample

Sample size was estimated for a paired t-test using a priori predicted effect size of 0.48 (Park et al., 2006), and 38 consecutive participants were enrolled (alpha=0.05, power=0.80, +15% attrition) after obtaining informed consent. Participants were included if they had a primary DSM-5 diagnosis of OCD (300.3), and a score ≥38.5 on self-reported cognitive difficulties assessed by CAIOC-13, were aged 18–50 years, on stable medication for at least two months, had a minimum 7 years of education, were fluent in English and selected Indian languages (Hindi/Bengali) and were right-handed with normal or corrected vision and hearing. Exclusion criteria were - comorbid psychotic disorders and bipolar disorders, substance dependence (except nicotine), behavioural addiction, high suicidal risk, neurological disorders, recent structured psychotherapy (≥6 sessions in the past 3 months), previous history of brain stimulation, or sensorimotor impairments interfering with participation.

### Tools

Assessments included standard screening, clinical, cognitive, personality, and neuropsychological measures. The Edinburgh Handedness Inventory–Short Form (Veale, 2014) and the Structured Clinical Interview for DSM-5 Disorders, Clinician Version (SCID-5-CV) (First et al., 2016) were administered at baseline (week 1). Clinical severity measures, including the Clinical Global Impression Scale (CGI) (Guy, 1976), the YBOCS, HAM-A and MADRS were administered at baseline, midline (weeks 3, 5, and 7), post-intervention (weeks 9–12), and follow-up (weeks 20–26). Self-report tools—the CAIOC-13, MARS, SDS and the Personality Inventory for DSM-5—Brief Form (PID-5-BF) (Krueger et al., 2012) were administered at designated intervals (baseline and post-intervention for all, except the PID-5-BF, which was administered only at baseline; CAIOC-13 was additionally administered at midline and follow-up). All neuropsychological assessments were conducted at baseline and post-intervention, with the SST additionally administered at follow-up. Alternate equivalent forms of neuropsychological tests were used at post-intervention to minimize practice effects.

### Procedure

The intervention was first adapted and validated through a systematic process that included review of the existing program, selection of suitable cognitive training tasks, development of a new training task for interference control (IC); expert validation, incorporation of feedback, and preparation of standard operating procedures and fidelity checklists and a preliminary trial on three participants. Task validation was carried out both with clinicians and with clients, the latter limited to the new IC task, as other existing tasks had previously been validated in six patients in earlier studies (Kashyap et al., 2019; Rini et al., 2023). The adapted intervention was then evaluated in an open-label efficacy trial involving 38 patients with OCD (Fig. 1). Data collection occurred between November 2020 and January 2023. Data collection and participant screening took place from November 2020 to January 2023. Following the COVID-19 outbreak, the study protocol was amended for remote delivery with Institute Ethics Committee (IEC) approval, including digital consent, online videoconferencing sessions, and task adaptation using curated mobile apps and online forms. Data consistency and security complied with IEC guidelines.

**Figure 1.**
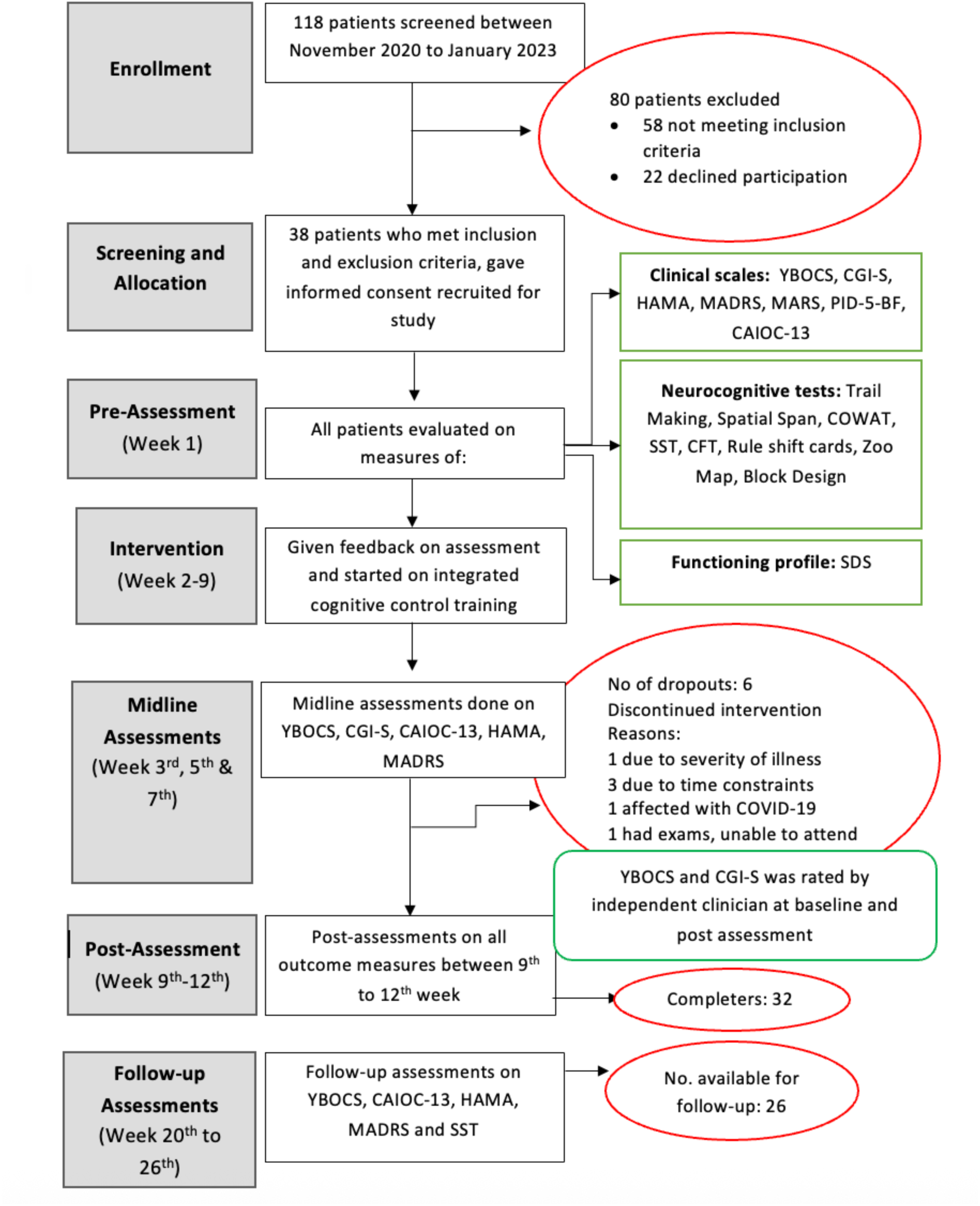
Flowchart depicting the recruitment process in Phase 2. YBOCS *=* Yale–Brown Obsessive–Compulsive Scale; *CGI-S* = Clinical Global Impression–Severity Scale; *HAMA* = Hamilton Anxiety Rating Scale; *MADRS* = Montgomery–Åsberg Depression Rating Scale; *PID-5BF* = Personality Inventory for DSM-5–Brief Form; *CAIOC-13* = Cognitive Assessment Instrument for Obsessions and Compulsions–13 Item Version; *COWAT* = Controlled Oral Word Association Test; *SST* = Stop Signal Task; *CFT* = Rey–Osterrieth Complex Figure Test; *SDS* = Sheehan Disability Scale.

### Intervention

The Integrated Cognitive Control Training Program (ICCT) was expanded and modified from the two prior studies (Kashyap et al., 2019; Rini et al., 2023). ICCT comprised 24 hours of training over eight weeks. Each week featured a one-hour therapist-led session, along with two hours of homework divided into four 20-30-minute sessions. The program consisted of three main components (Table 1): cognitive activation, metacognitive strategy monitoring, and generalization (Bowie et al., 2013). Cognitive activation used progressively challenging smartphone-based tasks like Cogtrain, Marbel Solitaire, Four in a Row, Sudoku, and the developed interference control tasks, aimed at enhancing neuroplasticity (B. A. Wilson, 2008). Strategy Monitoring focused on metacognitive processes through games, mindful colouring, and meditation. Generalization promoted real-life application through role-plays, simulations, and real-world practice. Homework sessions involved smartphone-based cognitive tasks, journaling, and real-life exercises for skill generalization.

**Table 1:**
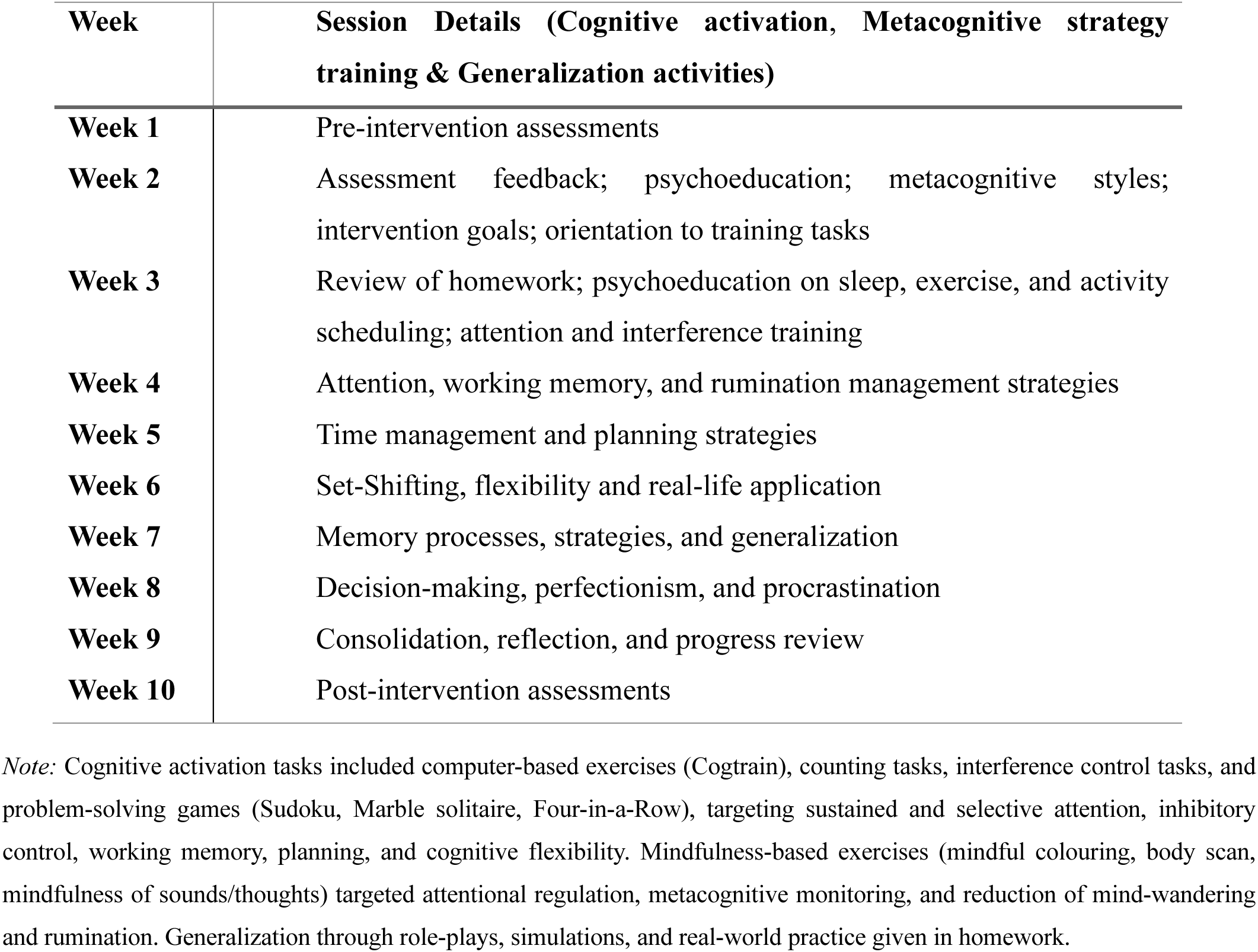
Session details & schedule of activities of Integrated Cognitive Control Training Program (ICCT).

| Week | Session Details (Cognitive activation, Metacognitive strategy training & Generalization activities) |
| --- | --- |
| Week 1 | Pre-intervention assessments |
| Week 2 | Assessment feedback; psychoeducation; metacognitive styles; intervention goals; orientation to training tasks |
| Week 3 | Review of homework; psychoeducation on sleep, exercise, and activity scheduling; attention and interference training |
| Week 4 | Attention, working memory, and rumination management strategies |
| Week 5 | Time management and planning strategies |
| Week 6 | Set-Shifting, flexibility and real-life application |
| Week 7 | Memory processes, strategies, and generalization |
| Week 8 | Decision-making, perfectionism, and procrastination |
| Week 9 | Consolidation, reflection, and progress review |
| Week 10 | Post-intervention assessments |

*Note:* Cognitive activation tasks included computer-based exercises (Cogtrain), counting tasks, interference control tasks, and problem-solving games (Sudoku, Marble solitaire, Four-in-a-Row), targeting sustained and selective attention, inhibitory control, working memory, planning, and cognitive flexibility. Mindfulness-based exercises (mindful colouring, body scan, mindfulness of sounds/thoughts) targeted attentional regulation, metacognitive monitoring, and reduction of mind-wandering and rumination. Generalization through role-plays, simulations, and real-world practice given in homework.

### Analysis

Statistical analyses were performed using RStudio (version 3.3.0+). Descriptive statistics summarized clinical and sociodemographic variables. Intervention effects across pre-, post-, and follow-up assessments were examined using linear mixed-effects models. Follow-up analyses of primary outcomes (CAIOC-13, SST) and key clinical measures (YBOCS, HAM-A, MADRS) included 26 participants and were conducted using maximum likelihood estimation, incorporating all available data under a missing-at-random assumption (pre n = 38, post n = 35, follow-up n = 26).

## Results

The sociodemographic and clinical characteristics of the sample are presented in Table 2. Out of the 38 patients, 36 patients owned a smartphone and were able to access the online platform; the other two took help from family (husband/mother). The average age of onset of OCD in the sample was around 19 years (± 5.46 years), with a mean illness duration of about 10 years (± 5.46 years). All patients included were on outpatient treatment with a stable dose of medication (SSRIs) for the past two months.

**Table 2:**
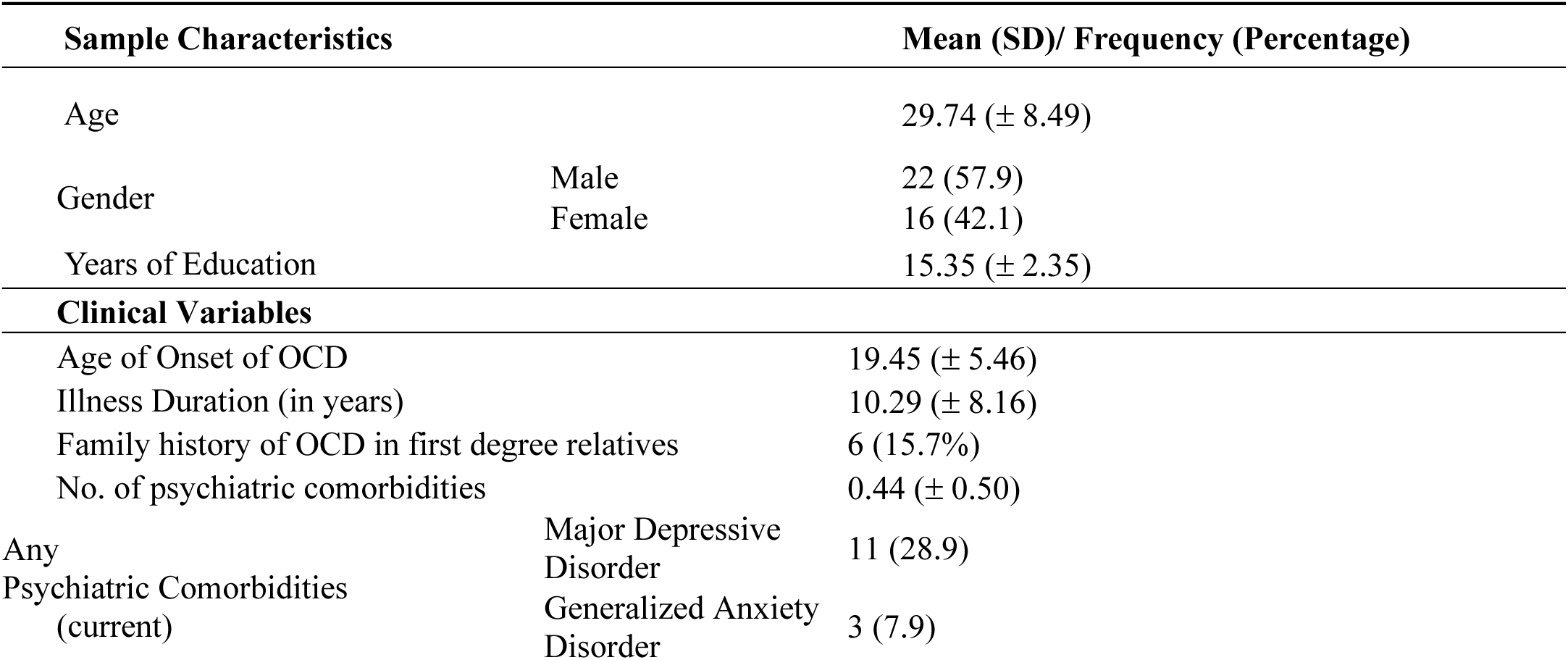
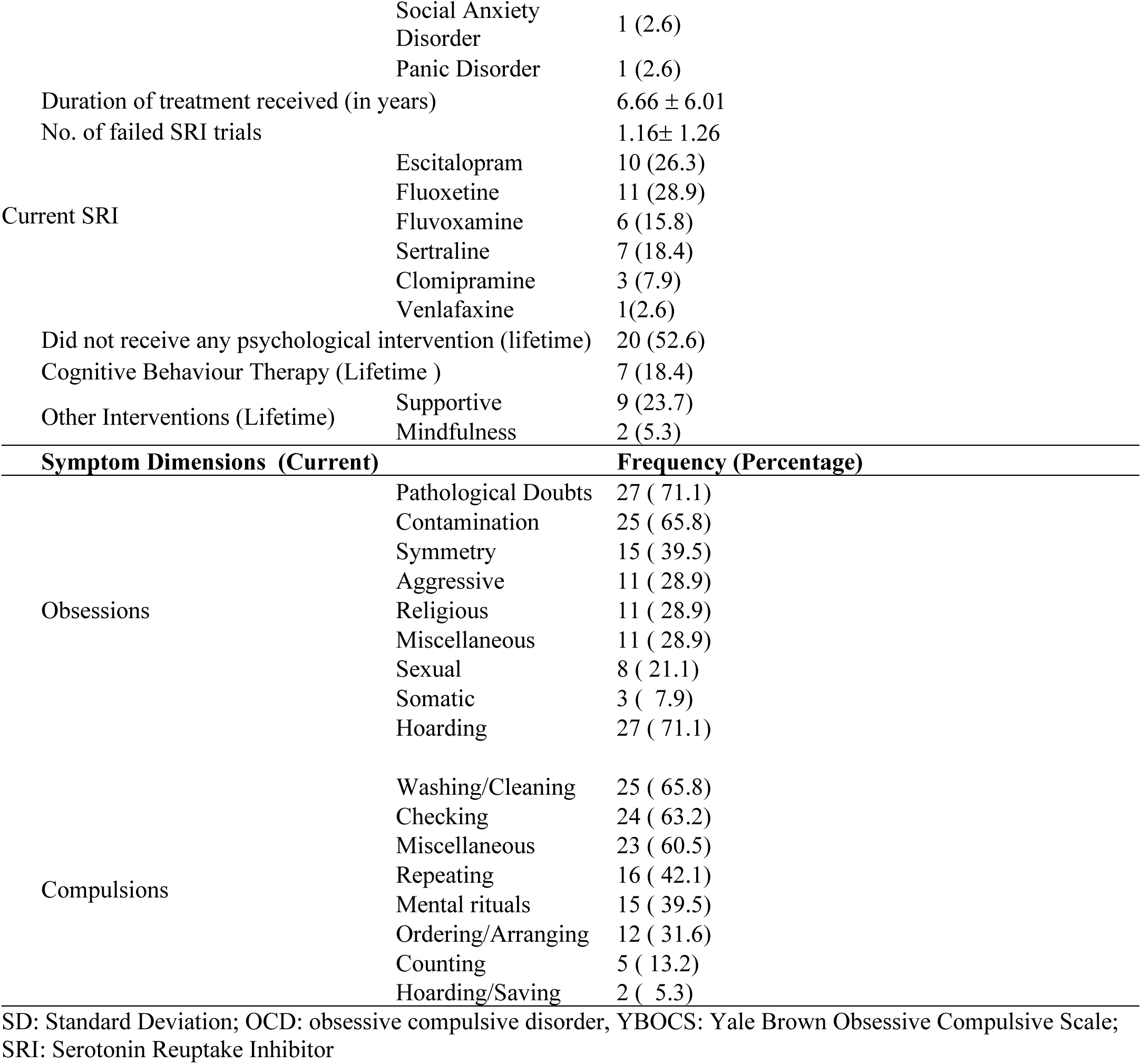
Sociodemographic and clinical characteristics (N=38).

Table 3 shows that the treatment had a significant impact on reducing cognitive difficulties reported by patients (CAIOC-13). However, the effect on response inhibition, as measured by the stop signal reaction time, was minimal. Notably, all clinical scales showed significant improvement, with a strong reduction in OCD severity, anxiety, and depression scores. At 3 months follow-up, there was significant improvement on most measures, except on SSRT. From table 3, it is seen that most variables had large improvements at 3 months follow-up, except SSRT (moderate effect).

**Table 3:** Fixed-Effect Estimates with Marginal R² from Linear Mixed-Effects Models for Primary, Secondary, and Neuropsychological Outcome Variables.

| Measure | Pre (M ± SD) (n=38) | Post (M ± SD) (n=35) | Follow-up (M ± SD) (n=26) | β | SE | 95% CI | t(df) | R²(M) |
| --- | --- | --- | --- | --- | --- | --- | --- | --- |
| Primary Outcome Variables |  |  |  |  |  |  |  |  |
| CAIOC-13 | 48.79 ± 6.99 | 30.60 ± 10.36 | 23.75 ± 13.42 | −18.19 | 2.94 | [−24.13, −12.25] | −6.19 (49) | .58 |
| SSRT (ms) | 254.00 ± 58.41 | 232.00 ± 44.92 | 223.74 ± 40.14 | −12.44 | 5.76 | [−23.99, −0.89] | −2.16 (49) | .11 |
| Secondary Outcome Variables |  |  |  |  |  |  |  |  |
| Y-BOCS Total | 23.87 ± 4.97 | 17.77 ± 6.07 | 7.83 ± 6.23 | −7.84 | 1.38 | [−10.61, −5.07] | −5.68 (49) | .54 |
| HAM-A | 13.24 ± 4.88 | 5.23 ± 3.03 | 9.83 ± 5.10 | −8.00 | 1.31 | [−10.63, −5.37] | −6.12 (49) | .61 |
| MADRS | 19.87 ± 7.36 | 8.51 ± 4.30 | 14.67 ± 8.47 | −10.91 | 1.79 | [−14.52, −7.30] | −6.09 (49) | .60 |
| CGI-S | 4.71 ± 0.65 | 3.83 ± 0.92 | 1.13 ± 0.80 | −1.12 | 0.21 | [−1.55, −0.69] | −5.33 (49) | .50 |
| MARS | 65.68 ± 10.91 | 79.06 ± 10.10 | — | 13.38 | 2.89 | [7.56, 19.20] | 4.63 (49) | .30 |
| SDS-Work | 6.32 ± 1.83 | 4.03 ± 2.08 | — | −2.20 | 0.53 | [−3.27, −1.13] | −4.17 (49) | .26 |
| SDS-Family | 5.87 ± 2.23 | 3.88 ± 2.18 | — | −1.94 | 0.61 | [−3.17, −0.71] | −3.18 (49) | .18 |
| Neuropsychological Measures |  |  |  |  |  |  |  |  |
| TMT-A (s) | 44.36 ±<br>14.04 | 37.32 ±<br>13.17 | — | −6.84 | 2.41 | [−<br>11.68, −<br>2.00] | −2.84<br>(49) | .14 |
| TMT-B (s) | 108.95 ±<br>49.93 | 84.13 ±<br>34.29 | — | −<br>21.09 | 7.38 | [−<br>35.92, −<br>6.26] | −2.86<br>(49) | .15 |
| Spatial Span<br>Total | 13.49 ±<br>2.46 | 16.29 ±<br>2.07 | — | 2.18 | 0.58 | [1.02,<br>3.34] | 3.76<br>(49) | .23 |
| COWAT | 11.70 ±<br>3.29 | 14.33 ±<br>3.19 | — | 2.49 | 0.67 | [1.14,<br>3.84] | 3.72<br>(49) | .22 |
| CFT<br>Immediate<br>Recall | 35.78 ±<br>7.41 | 35.97 ±<br>4.65 | — | 5.89 | 1.56 | [2.77,<br>9.01] | 3.77<br>(49) | .23 |
| CFT<br>Delayed<br>Recall | 23.14 ±<br>7.30 | 28.74 ±<br>4.50 | — | 6.97 | 1.64 | [3.68,<br>10.26] | 4.26<br>(49) | .26 |
| Rule-Shift<br>Profile | 2.86 ±<br>0.67 | 3.64 ±<br>0.49 | — | 0.69 | 0.18 | [0.32,<br>1.06] | 3.83<br>(49) | .24 |
| Zoo-Map<br>Profile | 2.38 ±<br>1.06 | 3.39 ±<br>0.66 | — | 0.91 | 0.21 | [0.48,<br>1.34] | 4.36<br>(49) | .27 |

Note. Values are means ± standard deviations. $\beta$ = unstandardized fixed-effect estimate for change across time; SE = standard error; CI = confidence interval; $t(df)$ = Satterthwaite-approximated test of fixed effect; $R^2(M)$ = marginal $R^2$ (variance explained by fixed effects). CAIOC-13 = Cognitive Assessment Instrument for Obsessions and Compulsions; SSRT = Stop-Signal Reaction Time; Y-BOCS = Yale–Brown Obsessive–Compulsive Scale; HAM-A = Hamilton Anxiety Rating Scale; MADRS = Montgomery–Åsberg Depression Rating Scale; CGI-S = Clinical Global Impression–Severity; MARS = Metacognitive Awareness and Regulation Scale; SDS = Sheehan Disability Scale; TMT = Trail Making Test; COWAT = Controlled Oral Word Association Test; CFT = Complex Figure Task.

### Acceptability and feasibility outcomes of the ICCT Program

Overall satisfaction with the ICCT program was high, with 71% of participants reporting being very satisfied; 77% rated the intervention as very useful for managing cognitive difficulties and 23% as somewhat useful, with similar ratings for symptom management and future use. Treatment fidelity was well maintained. Of the 304 ICCT sessions delivered, 60.5% were conducted exclusively using ICCT procedures, with no additional techniques. Non-ICCT techniques were used infrequently, most often limited to brief supportive psychoeducation

(13.1%). Other techniques (arousal reduction, CBT strategies, and family interventions) were used only occasionally (each ≤5.2%, typically in one to three sessions). Participant responsiveness was reflected in homework completion (mean = 10.578 ± 3.789 of 16 hours) and task engagement, with 78% reaching the ‘medium’ level by session 4 and 60% the ‘hard’ level by session 8. Dropout analyses showed that non-completers had longer illness duration (U = 41, p = 0.029), more psychiatric comorbidities (U = 44.5, p = 0.024), longer treatment history (U = 27.5, p = 0.007), and higher baseline OCD severity (YBOCS; U = 36.5, p = 0.018) compared to completers.

## Discussion

The present study aimed to examine the effectiveness of an 8-week integrated cognitive control training (ICCT) program on cognitive functioning in individuals with OCD and to explore potential correlates of change in neuropsychological, clinical, and socio-occupational outcomes. The main findings were significant improvement in self-perceived cognitive difficulties, along with improvement in other neuropsychological functions and improvement in symptom severity and functioning.

### Efficacy on primary outcome measures

Following ICCT, a large improvement was observed on CAOIC, post-intervention, with 71% of participants scoring below the impairment cut-off, indicating substantial reductions in perceived cognitive difficulties, especially in everyday functional domains such as decision-making, procrastination, and perfectionism. Participant feedback also reflected the perceived benefits of the ICCT program. Subjective measures of cognition, as part of a comprehensive neuropsychological assessment, reflect real-life impairment (Rabin et al., 2015; Van Patten et al., 2025), though prior research often lacked subjective outcome measures (Haug et al., 2013).

Treatment effect was found to be small on most of the response inhibition functions [e.g., stop signal reaction time (R²₍M₎ = .11)], which indicates that the ICCT intervention was less effective in improving response inhibition deficits. A change (decrease) of only 19 milliseconds was noted in the Stop Signal Reaction Time (SSRT) in the post-assessment. The limited effect on response inhibition may be attributable to trait-like vulnerabilities (Chamberlain et al., 2006, 2007; Menzies et al., 2007), lack of clarity regarding the deficit in OCD, i.e., response inhibition versus heightened error monitoring (Berlin & Lee, 2018; Kalanthroff et al., 2017; Silveira et al., 2020), or inadequate dose (duration of training) effects, as studies which have showed improvements in response inhibition had had 24–25 sessions of training (Johnstone et al., 2012; ten Brinke et al., 2020). Additionally, several studies used variations of the Go/No-Go or SST task for training response inhibition which is a bottom-up approach resulting in near transfer (You et al., 2023). Some researchers have concluded that top-down controlled response inhibition is difficult or impossible to improve (Enge et al., 2014; You et al., 2023).

### Efficacy on secondary outcome measures

Moderate treatment effects were observed in non-verbal memory, set shifting, cognitive flexibility, planning, visuospatial working memory and verbal fluency, and in sustained attention. Most of these findings are consistent with prior research on cognitive training in OCD (Cameron et al., 2020; Kashyap et al., 2019; Rini et al., 2023; van Passel et al., 2020), other psychiatric and neurological disorders (Anderson et al., 2021; Jennings et al., 2005; Kurtz, 2012; Motter et al., 2019), and aging populations (Adam et al., 2017; Szöke et al., 2008), though some studies have reported mixed or null findings (Grönholm-Nyman et al., 2017; Rodewald et al., 2011; Solari et al., 2004).

The cognitive-training program yielded a large effect-size reduction in OCD severity, with 47% of participants classified as responders (>35% reduction on the YBOCS) (Mataix-Cols et al., 2016). The goal of cognitive training is to facilitate lasting improvement in cognitive functions themselves, and to generalize improvements to other domains (Keshavan et al., 2014; B. A. Wilson, 2008). Most prior cognitive training trials in OCD showed limited clinical transfer (Buhlmann et al., 2015; Cameron et al., 2020), though some reported symptom improvements (Kashyap et al., 2019; Rini et al., 2023). Improvement on the Y-BOCS, suggests a potential transfer of benefit comparable in magnitude to response rates typically observed with CBT or pharmacotherapy (Foa, 2010), though this should be interpreted cautiously given the absence of a control group.

The ICCT intervention significantly improved metacognitive awareness and reduced socio-occupational disability across work, social, and family domains, aligning with prior findings in OCD and other psychiatric disorders (Bowie et al., 2013; Kashyap et al., 2019; Montemagni et al., 2021; Rini et al., 2023). Metacognitive strategy monitoring, also called performance monitoring, was incorporated in the intervention to help individuals build new strategies, recognize which strategies might work given the situation, and develop skills to flexibly switch strategies as an adaptation to shifting environmental demands (Bowie et al., 2013).

The improvements in the above cognitive functions may have led to better attentional control, helping individuals manage intrusive thoughts by redirecting focus to relevant tasks, thereby reducing the salience of obsessions (Salkovskis, 2007). Enhancing visuospatial working memory may reduce uncertainty and checking behaviours (Jaafari et al., 2013; Lambrecq et al., 2014) and has been associated with reduced anxiety and compulsive symptoms (Wang et al., 2022). Gains in cognitive flexibility and set-shifting may lead to improved switching abilities and verbal fluency, helping reduce perseverative behaviour (Y. S. Shin et al., 2012). Additionally, improved planning skills support better management of daily life and behavioural change (Martoni et al., 2018), while enhanced organizational strategies may help individuals adopt more adaptive problem-solving approaches and reduce symptom focus (Meyer & Meyer, 1995; Shorr et al., 1992).

In the current study, participants continued to improve and maintained gains even at 3-month follow-up in terms of symptom severity and cognitive functioning in daily life. Because the treatment factors were not kept stable during the follow-up phase, this should be regarded with caution. Nevertheless, the maintenance at follow-up is also reflected by the fact that most individuals were able to resume their expected socio-occupational roles without interference from their previous OCD-related functional deficits.

### Acceptability and Feasibility of the program

The ICCT program demonstrated strong acceptability and feasibility, aligning with previous findings in OCD and cognitive intervention research (Cameron et al., 2020). High participant satisfaction and engagement suggest that participants appreciated interactive tasks and reduced anxiety due to non-direct symptom targeting. Demand was reflected in a low dropout rate comparable to Passel et al. (2020), with dropouts showing greater illness severity and comorbidity, consistent with Aderka et al. (2011). Implementation was supported through a blended delivery mode (Wentzel et al., 2016), and high treatment fidelity was ensured by minimizing non-study techniques, reinforcing internal validity (Fonagy & Luyten, 2019; Onken et al., 2014). Participant responsiveness was evidenced by good homework compliance and active task engagement, with progressive improvement across tasks targeting working memory, attention, planning, and set-shifting. These findings highlight the program’s feasibility and effectiveness in enhancing cognitive functioning in OCD (Bowen et al., 2009).

The present study has several strengths, including a clinically representative and homogeneous sample, strict inclusion criteria, comprehensive assessments, and participants maintained on a stable medication regimen without alterations during the trial to minimize confounding effects. The structured, evidence-based ICCT intervention used validated tasks, ensured treatment fidelity, monitored adherence, and applied blind ratings and intent-to-treat analysis. It was designed in accordance with evidence-based recommendations for achieving far transfer effects of cognitive training. The specific components of the intervention study like cognitive stimulation, metacognitive training, which were incorporated through mindfulness activities and reflective exercises after each trial of cognitive stimulation tasks may have helped in better management of obsessions, anxiety and depressive symptoms that the OCD individuals were undergoing. Low dropout rates, post-intervention feedback, and follow-up assessment supported its feasibility and acceptability. However, limitations include its open-label design, lack of process variable assessments, and limited benefits for individuals with very severe OCD. Clinically, ICCT shows promise in addressing cognitive and functional deficits in OCD, particularly as an adjunct to SSRIs or CBT, and is well suited for scalable implementation in resource-limited settings, as its blended delivery model and digitized, task-based format allow for adaptation to online and remote platforms. ICCT warrants evaluation in larger randomized controlled trials and may be applied to other disorders characterized by cognitive control deficits. Finally, it emphasizes the training value of manualizing and disseminating ICCT as a gamified, technology-driven tool in clinical and research contexts.

### Conclusion

The current study’s findings indicate the potential of using cognitive training programs in addition to traditional CBT interventions to achieve global improvements in OCD. Future studies will need to replicate and strengthen the results using a control arm and randomized designs, along with multimodal assessments including neuroimaging to compare different interventions with ICCT and derive stronger conclusions.

## Data Availability

All data produced in the present study are available upon reasonable request to the authors

## References

Abramovitch, A., Abramowitz, J. S., & Mittelman, A. (2013). The neuropsychology of adult obsessive–compulsive disorder: A meta-analysis. Clinical Psychology Review, 33(8), 1163–1171. 10.1016/j.cpr.2013.09.004

Adam, K. C. S., Vogel, E. K., & Awh, E. (2017). Clear evidence for item limits in visual working memory. Cognitive Psychology, 97, 79–97. 10.1016/j.cogpsych.2017.07.001

Anderson, A. C., Youssef, G. J., Robinson, A. H., Lubman, D. I., & Verdejo-Garcia, A. (2021). Cognitive boosting interventions for impulsivity in addiction: A systematic review and meta-analysis of cognitive training, remediation and pharmacological enhancement. Addiction (Abingdon, England), 116(12), 3304–3319. 10.1111/add.15469

Bannon, S., Gonsalvez, C. J., Croft, R. J., & Boyce, P. M. (2006). Executive functions in obsessive-compulsive disorder: State or trait deficits? The Australian and New Zealand Journal of Psychiatry, 40(11–12), 1031–1038. 10.1080/j.1440-1614.2006.01928.x

Benton, A., Hamsher, K. deS, & Sivan, A. (1976). Multilingual Aphasia Examination. Iowa City. University of Iowa.

Berlin, G. S., & Lee, H.-J. (2018). Response inhibition and error-monitoring processes in individuals with obsessive-compulsive disorder. Journal of Obsessive-Compulsive and Related Disorders, 16, 21–27. 10.1016/j.jocrd.2017.11.001

Bhattacharya, M., Kashyap, H., & Reddy, Y. C. J. (2024). Cognitive Training in Obsessive-Compulsive Disorder: A Systematic Review. Indian Journal of Psychological Medicine, 46(2), 110–118. 10.1177/02537176231207781

Bowen, D. J., Kreuter, M., Spring, B., Cofta-Woerpel, L., Linnan, L., Weiner, D., Bakken, S., Kaplan, C. P., Squiers, L., Fabrizio, C., & Fernandez, M. (2009). How We Design Feasibility Studies. American Journal of Preventive Medicine, 36(5), 452–457. 10.1016/j.amepre.2009.02.002

Bowie, C. R. (2019). Cognitive remediation for severe mental illness: State of the field and future directions. World Psychiatry, 18(3), 274–275. 10.1002/wps.20660

Bowie, C. R., Gupta, M., Holshausen, K., Jokic, R., Best, M., & Milev, R. (2013). Cognitive remediation for treatment-resistant depression: Effects on cognition and functioning and the role of online homework. The Journal of Nervous and Mental Disease, 201(8), 680–685. 10.1097/NMD.0b013e31829c5030

Bowie, C. R., & Harvey, P. D. (2006). Administration and interpretation of the Trail Making Test. Nature Protocols, 1(5), 2277–2281.

Buhlmann, U., Wacker, R., & Dziobek, I. (2015). Inferring other people’s states of mind: Comparison across social anxiety, body dysmorphic, and obsessive-compulsive disorders. Journal of Anxiety Disorders, 34, 107–113. 10.1016/j.janxdis.2015.06.003

Calkins, A. W., & Otto, M. W. (2013). Testing the Boundaries of Computerized Cognitive Control Training on Symptoms of Obsessive Compulsive Disorder. Cognitive Therapy and Research, 37(3), 587–594. 10.1007/s10608-012-9496-x

Cameron, D. H., McCabe, R. E., Rowa, K., O’Connor, C., & McKinnon, M. C. (2020). A pilot study examining the use of Goal Management Training in individuals with obsessive-compulsive disorder. Pilot and Feasibility Studies, 6, 151. 10.1186/s40814-020-00684-0

Cavedini, P., Zorzi, C., Piccinni, M., Cavallini, M. C., & Bellodi, L. (2010). Executive dysfunctions in obsessive-compulsive patients and unaffected relatives: Searching for a new intermediate phenotype. Biological Psychiatry, 67(12), 1178–1184. 10.1016/j.biopsych.2010.02.012

Chamberlain, S. R., Fineberg, N. A., Blackwell, A. D., Robbins, T. W., & Sahakian, B. J. (2006). Motor inhibition and cognitive flexibility in obsessive-compulsive disorder and trichotillomania. The American Journal of Psychiatry, 163(7), 1282–1284. 10.1176/appi.ajp.163.7.1282

Chamberlain, S. R., Fineberg, N. A., Menzies, L. A., Blackwell, A. D., Bullmore, E. T., Robbins, T. W., & Sahakian, B. J. (2007). Impaired Cognitive Flexibility and Motor Inhibition in Unaffected First-Degree Relatives of Patients With Obsessive-Compulsive Disorder. American Journal of Psychiatry, 164(2), 335–338. 10.1176/ajp.2007.164.2.335

Cicerone, K. D., Langenbahn, D. M., Braden, C., Malec, J. F., Kalmar, K., Fraas, M., Felicetti, T., Laatsch, L., Harley, J. P., Bergquist, T., Azulay, J., Cantor, J., & Ashman, T. (2011). Evidence-based cognitive rehabilitation: Updated review of the literature from 2003 through 2008. Archives of Physical Medicine and Rehabilitation, 92(4), 519–530. 10.1016/j.apmr.2010.11.015

D’Alcante, C. C., Diniz, J. B., Fossaluza, V., Batistuzzo, M. C., Lopes, A. C., Shavitt, R. G., Deckersbach, T., Malloy-Diniz, L., Miguel, E. C., & Hoexter, M. Q. (2012). Neuropsychological predictors of response to randomized treatment in obsessive-compulsive disorder. Progress in Neuro-Psychopharmacology & Biological Psychiatry, 39(2), 310–317. 10.1016/j.pnpbp.2012.07.002

Dittrich, W. H., Johansen, T., & Fineberg, N. A. (2011). Cognitive Assessment Instrument of Obsessions and Compulsions (CAIOC-13)—A new 13-item scale for evaluating functional impairment associated with OCD. Psychiatry Research, 187(1–2), 283–290. 10.1016/j.psychres.2010.10.031

Enge, S., Behnke, A., Fleischhauer, M., Küttler, L., Kliegel, M., & Strobel, A. (2014). No evidence for true training and transfer effects after inhibitory control training in young healthy adults. Journal of Experimental Psychology: Learning, Memory, and Cognition, 40(4), 987.

First, M. B., Williams, J. B. W., Karg, R. S., & Spitzer, R. L. (2016). User’s guide for the SCID-5-CV Structured Clinical Interview for DSM-5® disorders: Clinical version (pp. xii, 158). American Psychiatric Publishing, Inc.

Foa, E. B. (2010). Cognitive behavioral therapy of obsessive-compulsive disorder. Dialogues in Clinical Neuroscience, 12(2), 199–207. https://www.ncbi.nlm.nih.gov/pmc/articles/PMC3181959/

Fonagy, P., & Luyten, P. (2019). Fidelity vs. flexibility in the implementation of psychotherapies: Time to move on. World Psychiatry, 18(3), 270–271. 10.1002/wps.20657

Fruehauf, L. M., Fair, J. E., Liebel, S. W., Bjornn, D., & Larson, M. J. (2021). Cognitive control in obsessive-compulsive disorder (OCD): Proactive control adjustments or consistent performance? Psychiatry Research, 298, 113809. 10.1016/j.psychres.2021.113809

Goodman, W. K., Price, L. H., Rasmussen, S. A., Mazure, C., Fleischmann, R. L., Hill, C. L., Heninger, G. R., & Charney, D. S. (1989). The Yale-Brown Obsessive Compulsive Scale. I. Development, use, and reliability. Archives of General Psychiatry, 46(11), 1006–1011. 10.1001/archpsyc.1989.01810110048007

Grönholm-Nyman, P., Soveri, A., Rinne, J. O., Ek, E., Nyholm, A., Stigsdotter Neely, A., & Laine, M. (2017). Limited Effects of Set Shifting Training in Healthy Older Adults. Frontiers in Aging Neuroscience, 9. https://www.frontiersin.org/articles/10.3389/fnagi.2017.00069

Gurappa, P. (2009). Wechsler Memory Scale.

Guy, W. (1976). Clinical global impressions scale. Psychiatry.

Hamilton, M. (1959). The Assessment of Anxiety States by Rating. British Journal of Medical Psychology, 32(1), 50–55. 10.1111/j.2044-8341.1959.tb00467.x

Haug, E. T., Havnen, A., Hansen, B., Bless, J., & Kvale, G. (2013). Attention training with dichotic listening in OCD patients using an iPhone/iPod app. Clinical Neuropsychiatry: Journal of Treatment Evaluation, 10(3, Suppl 1), 45–47. https://www.proquest.com/docview/1450173726/C89483DE966E4139PQ/25

Hybel, K. A., Mortensen, E. L., Lambek, R., Højgaard, D. R. M. A., & Thomsen, P. H. (2017). Executive function predicts cognitive-behavioral therapy response in childhood obsessive-compulsive disorder. Behaviour Research and Therapy, 99, 11–18. 10.1016/j.brat.2017.08.009

Jaafari, N., Frasca, M., Rigalleau, F., Rachid, F., Gil, R., Olié, J.-P., Guehl, D., Burbaud, P., Aouizerate, B., & Rotgé, J.-Y. (2013). Forgetting what you have checked: A link between working memory impairment and checking behaviors in obsessive-compulsive disorder. European Psychiatry, 28(2), 87–93.

Jennings, J. M., Webster, L. M., Kleykamp, B. A., & Dagenbach, D. (2005). Recollection Training and Transfer Effects in Older Adults: Successful Use of a Repetition-Lag Procedure. Aging, Neuropsychology, and Cognition, 12(3), 278–298. 10.1080/138255890968312

Johnstone, S. J., Roodenrys, S., Blackman, R., Johnston, E., Loveday, K., Mantz, S., & Barratt, M. F. (2012). Neurocognitive training for children with and without AD/HD. ADHD Attention Deficit and Hyperactivity Disorders, 4, 11–23.

Kalanthroff, E., Henik, A., Simpson, H. B., Todder, D., & Anholt, G. E. (2017). To Do or Not to Do? Task Control Deficit in Obsessive-Compulsive Disorder. Behavior Therapy, 48(5), 603–613. 10.1016/j.beth.2017.01.004

Karbach, J., & Kray, J. (2009). How useful is executive control training? Age differences in near and far transfer of task-switching training. Developmental Science, 12(6), 978–990. 10.1111/j.1467-7687.2009.00846.x

Kashyap, H., Kumar, J. K., Kandavel, T., & Reddy, Y. C. J. (2013). Neuropsychological functioning in obsessive-compulsive disorder: Are executive functions the key deficit? Comprehensive Psychiatry, 54(5), 533–540. 10.1016/j.comppsych.2012.12.003

Kashyap, H., Reddy, P., Mandadi, S., Narayanaswamy, J. C., Sudhir, P. M., & Reddy, Y. C. J. (2019). Cognitive training for neurocognitive and functional impairments in obsessive compulsive disorder: A case report. Journal of Obsessive-Compulsive and Related Disorders, 23, 7. APA PsycInfo®. 10.1016/j.jocrd.2019.100480

Keshavan, M. S., Vinogradov, S., Rumsey, J., Sherrill, J., & Wagner, A. (2014). Cognitive training in mental disorders: Update and future directions. The American Journal of Psychiatry, 171(5), 510–522. 10.1176/appi.ajp.2013.13081075

Kessler, R. C., Berglund, P., Demler, O., Jin, R., Merikangas, K. R., & Walters, E. E. (2005). Lifetime Prevalence and Age-of-Onset Distributions of DSM-IV Disorders in the National Comorbidity Survey Replication. Archives of General Psychiatry, 62(6), 593–602. 10.1001/archpsyc.62.6.593

Kim, M.-S., Park, S.-J., Shin, M. S., & Kwon, J. S. (2002). Neuropsychological profile in patients with obsessive-compulsive disorder over a period of 4-month treatment. Journal of Psychiatric Research, 36(4), 257–265.

Krueger, R. F., Derringer, J., Markon, K. E., Watson, D., & Skodol, A. E. (2012). Initial construction of a maladaptive personality trait model and inventory for DSM-5. Psychological Medicine, 42(9), 1879–1890. 10.1017/S0033291711002674

Kuelz, A. K., Hohagen, F., & Voderholzer, U. (2004). Neuropsychological performance in obsessive-compulsive disorder: A critical review. Biological Psychology, 65(3), 185–236. 10.1016/j.biopsycho.2003.07.007

Kurtz, M. M. (2012). Cognitive remediation for schizophrenia: Current status, biological correlates and predictors of response. Expert Review of Neurotherapeutics, 12(7), 813–821. 10.1586/ern.12.71

Lambrecq, V., Rotge, J.-Y., Jaafari, N., Aouizerate, B., Langbour, N., Bioulac, B., Liégeois-Chauvel, C., Burbaud, P., & Guehl, D. (2014). Differential role of visuospatial working memory in the propensity toward uncertainty in patients with obsessive–compulsive disorder and in healthy subjects. Psychological Medicine, 44(10), 2113–2124.

Lauenroth, A., Ioannidis, A. E., & Teichmann, B. (2016). Influence of combined physical and cognitive training on cognition: A systematic review. BMC Geriatrics, 16(1), 141. 10.1186/s12877-016-0315-1

Markarian, Y., Larson, M. J., Aldea, M. A., Baldwin, S. A., Good, D., Berkeljon, A., Murphy, T. K., Storch, E. A., & McKay, D. (2010). Multiple pathways to functional impairment in obsessive–compulsive disorder. Clinical Psychology Review, 30(1), 78–88. 10.1016/j.cpr.2009.09.005

Martoni, R. M., Risso, G., Giuliani, M., de Filippis, R., Cammino, S., Cavallini, C., & Bellodi, L. (2018). Evaluating Proactive Strategy in Patients With OCD During Stop Signal Task. Journal of the International Neuropsychological Society: JINS, 24(7), 703–714. Health & Medical Collection. 10.1017/S1355617718000267

Mataix-Cols, D., de la Cruz, L. F., Nordsletten, A. E., Lenhard, F., Isomura, K., & Simpson, H. B. (2016). Towards an international expert consensus for defining treatment response, remission, recovery and relapse in obsessive-compulsive disorder. World Psychiatry, 15(1), 80–81. 10.1002/wps.20299

McNamara, J. P. H., Reid, A. M., Balkhi, A. M., Bussing, R., Storch, E. A., Murphy, T. K., Graziano, P. A., Guzick, A. G., & Geffken, G. R. (2014). Self-Regulation and Other Executive Functions Relationship to Pediatric OCD Severity and Treatment Outcome. Journal of Psychopathology and Behavioral Assessment, 36(3), 432–442. 10.1007/s10862-014-9408-3

Menzies, L., Achard, S., Chamberlain, S., Fineberg, N., Chen, C., del Campo, N., & Bullmore, E. (2007). Neurocognitive endophenotypes of obsessive-compulsive disorder. Brain 130 (Pt 12), 3223–3236.

Meyer, J., & Meyer, K. (1995). The Meyers Scoring System for the Rey Complex Figure and the Recognition Trial. Psychological Assessment Resources, Odessa, FL.

Miyake, A., & Friedman, N. P. (2012). The Nature and Organization of Individual Differences in Executive Functions: Four General Conclusions. Current Directions in Psychological Science, 21(1), 8–14. 10.1177/0963721411429458

Montemagni, C., Del Favero, E., Riccardi, C., Canta, L., Toye, M., Zanalda, E., & Rocca, P. (2021). Effects of Cognitive Remediation on Cognition, Metacognition, and Social Cognition in Patients With Schizophrenia. Frontiers in Psychiatry, 12. https://www.frontiersin.org/articles/10.3389/fpsyt.2021.649737

Montgomery, S. A., & Asberg, M. (1979). A new depression scale designed to be sensitive to change. The British Journal of Psychiatry: The Journal of Mental Science, 134, 382–389. 10.1192/bjp.134.4.382

Motter, J. N., Grinberg, A., Lieberman, D. H., Iqnaibi, W. B., & Sneed, J. R. (2019). Computerized cognitive training in young adults with depressive symptoms: Effects on mood, cognition, and everyday functioning. Journal of Affective Disorders, 245, 28–37. 10.1016/j.jad.2018.10.109

Nielen, M. M. A., & Boer, J. a. D. (2003). Neuropsychological performance of OCD patients before and after treatment with fluoxetine: Evidence for persistent cognitive deficits. Psychological Medicine, 33(5), 917–925. 10.1017/S0033291703007682

Olley, A., Malhi, G., & Sachdev, P. (2007). Memory and executive functioning in obsessive–compulsive disorder: A selective review. Journal of Affective Disorders, 104(1), 15–23. 10.1016/j.jad.2007.02.023

Onken, L. S., Carroll, K. M., Shoham, V., Cuthbert, B. N., & Riddle, M. (2014). Reenvisioning clinical science: Unifying the discipline to improve the public health. Clinical Psychological Science, 2(1), 22–34.

Osterrieth, P. A. (1944). Le test de copie d’une figure complexe; contribution a l’etude de la perception et de la memoire. Archives de Psychologie.

Park, H. S., Shin, Y.-W., Ha, T. H., Shin, M. S., Kim, Y. Y., Lee, Y. H., & Kwon, J. S. (2006). Effect of cognitive training focusing on organizational strategies in patients with obsessive-compulsive disorder. Psychiatry and Clinical Neurosciences, 60(6), 718–726. 10.1111/j.1440-1819.2006.01587.x

Perna, G., Cavedini, P., Harvey, P. D., Di Chiaro, N. V., Daccò, S., & Caldirola, D. (2016). Does neuropsychological performance impact on real-life functional achievements in obsessive-compulsive disorder? A preliminary study. International Journal of Psychiatry in Clinical Practice, 20(4), 224–231. 10.1080/13651501.2016.1223856

Rabin, L. A., Smart, C. M., Crane, P. K., Amariglio, R. E., Berman, L. M., Boada, M., Buckley, R. F., Chételat, G., Dubois, B., Ellis, K. A., Gifford, K. A., Jefferson, A. L., Jessen, F., Katz, M. J., Lipton, R. B., Luck, T., Maruff, P., Mielke, M. M., Molinuevo, J. L.,…Sikkes, S. A. M. (2015). Subjective Cognitive Decline in Older Adults: An Overview of Self-Report Measures Used Across 19 International Research Studies. Journal of Alzheimer’s Disease: JAD, 48(0 1), S63–S86. 10.3233/JAD-150154

Rao, N. P., Reddy, Y. C. J., Kumar, K. J., Kandavel, T., & Chandrashekar, C. R. (2008). Are neuropsychological deficits trait markers in OCD? Progress in Neuro-Psychopharmacology & Biological Psychiatry, 32(6), 1574–1579. 10.1016/j.pnpbp.2008.05.026

Riesel, A., Endrass, T., Auerbach, L. A., & Kathmann, N. (2015). Overactive Performance Monitoring as an Endophenotype for Obsessive-Compulsive Disorder: Evidence From a Treatment Study. American Journal of Psychiatry, 172(7), 665–673. 10.1176/appi.ajp.2014.14070886

Rini, F., Kashyap, H., T, K., & Y.C., J. R. (2023). Cognitive Control Training Using a Novel Smartphone Application in Remitted Obsessive-Compulsive Disorder: A Pilot Study. Indian J Psychol Med, XX:1–3. 10.1177/02537176231172301

Rodewald, K., Rentrop, M., Holt, D. V., Roesch-Ely, D., Backenstraß, M., Funke, J., Weisbrod, M., & Kaiser, S. (2011). Planning and problem-solving training for patients with schizophrenia: A randomized controlled trial. BMC Psychiatry, 11(1), 73. 10.1186/1471-244X-11-73

Salkovskis, P. M. (2007). Psychological treatment of obsessive–compulsive disorder. Psychiatry, 6(6), 229–233. 10.1016/j.mppsy.2007.03.008

Sheehan, D. V., Mancini, M., Wang, J., Berggren, L., Cao, H., Dueñas, H. J., & Yue, L. (2016). Assessment of functional outcomes by Sheehan Disability Scale in patients with major depressive disorder treated with duloxetine versus selective serotonin reuptake inhibitors. Human Psychopharmacology, 31(1), 53–63. 10.1002/hup.2500

Shin, N. Y., Lee, T. Y., Kim, E., & Kwon, J. S. (2014). Cognitive functioning in obsessive-compulsive disorder: A meta-analysis. Psychological Medicine, 44(6), 1121–1130. 10.1017/S0033291713001803

Shin, Y. S., Shin, N. Y., Jang, J. H., Shim, G., Park, H. Y., Shin, M.-S., & Kwon, J. S. (2012). Switching strategy underlies phonemic verbal fluency impairment in obsessive–compulsive disorder. Journal of Obsessive-Compulsive and Related Disorders, 1(4), 221–227. 10.1016/j.jocrd.2012.07.005

Shorr, J. S., Delis, D. C., & Massman, P. J. (1992). Memory for the Rey-Osterrieth Figure: Perceptual clustering, encoding, and storage. Neuropsychology, 6(1), 43.

Silveira, V. P., Frydman, I., Fontenelle, L. F., Mattos, P., de Oliveira-Souza, R., Moll, J., Hoexter, M. Q., Miguel, E. C., McLaughlin, N. C. R., Shephard, E., & Batistuzzo, M. C. (2020). Exploring response inhibition and error monitoring in obsessive-compulsive disorder. Journal of Psychiatric Research, 126, 26–33. 10.1016/j.jpsychires.2020.04.002

Slagter, H. A., Davidson, R. J., & Lutz, A. (2011). Mental Training as a Tool in the Neuroscientific Study of Brain and Cognitive Plasticity. Frontiers in Human Neuroscience, 5, 17. 10.3389/fnhum.2011.00017

Snyder, H. R., Kaiser, R. H., Warren, S. L., & Heller, W. (2015). Obsessive-compulsive disorder is associated with broad impairments in executive function: A meta-analysis. Clinical Psychological Science: A Journal of the Association for Psychological Science, 3(2), 301–330. 10.1177/2167702614534210

Sofuoglu, M., DeVito, E. E., Waters, A. J., & Carroll, K. M. (2013). Cognitive enhancement as a treatment for drug addictions. Neuropharmacology, 64, 452–463. 10.1016/j.neuropharm.2012.06.021

Solari, A., Motta, A., Mendozzi, L., Pucci, E., Forni, M., Mancardi, G., & Pozzilli, C. (2004). Computer-aided retraining of memory and attention in people with multiple sclerosis: A randomized, double-blind controlled trial. Journal of the Neurological Sciences, 222(1), 99–104. 10.1016/j.jns.2004.04.027

Szöke, A., Trandafir, A., Dupont, M.-E., Méary, A., Schürhoff, F., & Leboyer, M. (2008). Longitudinal studies of cognition in schizophrenia: Meta-analysis. The British Journal of Psychiatry, 192(4), 248–257. 10.1192/bjp.bp.106.029009

Tate, R., Kennedy, M., Ponsford, J., Douglas, J., Velikonja, D., Bayley, M., & Stergiou-Kita, M. (2014). INCOG recommendations for management of cognition following traumatic brain injury, part III: Executive function and self-awareness. The Journal of Head Trauma Rehabilitation, 29(4), 338–352. 10.1097/HTR.0000000000000068

ten Brinke, L. F., Best, J. R., Chan, J. L. C., Ghag, C., Erickson, K. I., Handy, T. C., & Liu-Ambrose, T. (2020). The Effects of Computerized Cognitive Training With and Without Physical Exercise on Cognitive Function in Older Adults: An 8-Week Randomized Controlled Trial. The Journals of Gerontology: Series A, 75(4), 755–763. 10.1093/gerona/glz115

van Passel, B., Danner, U. N., Dingemans, A. E., Aarts, E., Sternheim, L. C., Becker, E. S., van Elburg, A. A., van Furth, E. F., Hendriks, G.-J., & Cath, D. C. (2020). Cognitive Remediation Therapy Does Not Enhance Treatment Effect in Obsessive-Compulsive Disorder and Anorexia Nervosa: A Randomized Controlled Trial. Psychotherapy and Psychosomatics, 89(4), 228–241. 10.1159/000505733

Van Patten, R., Mulhauser, K., Austin, T. A., Bellone, J. A., Cotton, E., Chan, L., Twamley, E. W., Sawyer, K., & LaFrance, W. C. (2025). The association between subjective and objective cognitive functioning from a transdiagnostic perspective: An umbrella review and meta-analysis. Clinical Psychology Review, 121, 102648. 10.1016/j.cpr.2025.102648

Veale, J. F. (2014). Edinburgh handedness inventory–short form: A revised version based on confirmatory factor analysis. Laterality: Asymmetries of Body, Brain and Cognition, 19(2), 164–177.

Verbruggen, F., & Logan, G. D. (2008). Response inhibition in the stop-signal paradigm. Trends in Cognitive Sciences, 12(11), 418–424. 10.1016/j.tics.2008.07.005

Vishwanathan, A., Kashyap, H., Reddy, R. P., Philip, M., Thippeswamy, H., & Desai, G. (2022). Neurocognition and Metacognition in Anxiety Disorders. Indian Journal of Psychological Medicine, 44(6), 558–566.

Wang, P., Yan, Z., Chen, T., Cao, W., Yang, X., Meng, F., Liu, Y., & Li, Z. (2022). Visuospatial working memory capacity moderates the relationship between anxiety and OCD related checking behaviors. Frontiers in Psychiatry, 13. 10.3389/fpsyt.2022.1039849

Wechsler, D. (1997). Wechsler Adult Intelligence Scale. The Psychological Corporation. San Antonio, TX.

Wentzel, J., Vaart, R. van der, Bohlmeijer, E. T., & Gemert-Pijnen, J. E. W. C. van. (2016). Mixing Online and Face-to-Face Therapy: How to Benefit From Blended Care in Mental Health Care. JMIR Mental Health, 3(1), e4534. 10.2196/mental.4534

Wilson, B. A. (2008). Neuropsychological rehabilitation. Annual Review of Clinical Psychology, 4, 141–162. 10.1146/annurev.clinpsy.4.022007.141212

Wilson, B., Alderman, N., Burgess, P., Emslie, H., & Evans, J. (1996). Behavioural assessment of the dysexecutive syndrome: Test manual. England: Thames Valley Test Company.

You, Y., Failla, A., & van der Kamp, J. (2023). No training effects of top-down controlled response inhibition by practicing on the stop-signal task. Acta Psychologica, 235, 103878. 10.1016/j.actpsy.2023.103878

